# The exposome and attention-related brain networks jointly predict attention problems in early adolescence

**DOI:** 10.64898/2026.03.26.26349404

**Authors:** Nia Berrian, Arielle S. Keller, Alfred F. Chao, Andrew J. Stier, Tyler M. Moore, Ran Barzilay, Marc G. Berman, Omid Kardan, Monica D. Rosenberg

**Affiliations:** Committee on Neurobiology, The University of Chicago, Chicago, IL; Medical Scientist Training Program, Pritzker School of Medicine, The University of Chicago, Chicago, IL; Institute for Mind and Biology, The University of Chicago, Chicago, IL; Department of Psychological Sciences, University of Connecticut, Storrs, CT; Institute for the Brain and Cognitive Sciences, University of Connecticut, Storrs, CT; Department of Psychology, The University of Chicago, Chicago, IL; The Santa Fe Institute, Santa Fe, NM; Department of Psychiatry, Perelman School of Medicine, University of Pennsylvania, Philadelphia, PA; Lifespan Brain Institute of Children’s Hospital of Pennsylvania and Penn Medicine, Philadelphia, PA; Department of Child and Adolescent Psychiatry and Behavioral Sciences, Children’s Hospital of Philadelphia, Philadelphia, PA; Neuroscience Institute, The University of Chicago, Chicago, IL; Department of Psychiatry, University of Michigan, Ann Arbor, MI

**Keywords:** Exposome, Attention, Connectome-based Predictive Modeling, Adolescence, ABCD Study

## Abstract

**Background:** Attention problems are common transdiagnostic symptoms of psychiatric illness. Although environmental exposures and experiences influence attention during adolescent development, the underlying neural pathways by which they do so is unclear.

**Methods:** We measured attention problems, attention-related brain networks, and multidimensional environmental experiences (the “exposome”) using data from the ABCD Study (N = 11,878). We tested whether the exposome is associated with 9-10-year-olds’ attention-related brain network strength and current and future attention problems. We further examined cross-sectional indirect pathways linking the exposome, brain network strength, and attention problems.

**Results:** The exposome predicted youths’ current and future self-, caregiver-, and teacher-reported attention problems as well as their current attention-related brain network strength. This brain network signature of sustained attention also predicted attention problems from all three reporters. Indirect effects models revealed that the exposome was associated with current reported attention problems both directly and indirectly though this brain signature. Conversely, predictive brain network strength was related to attention problems both directly and indirectly through the exposome.

**Conclusion:** Interactions between environmental exposures, experiences, and brain network organization are associated with attention problems in early adolescence. These findings support a bidirectional framework linking the environment and functional brain networks in the development of attention problems.

## Introduction

Development is shaped by dynamic, reciprocal interactions between an individual and their environment. For example, where a child lives affects where they go to school which in turn may affect their attention. At the same time, their attention function influences how they engage across multiple settings, shaping experiences in their academic, social, and everyday lives. Despite this reciprocity, most studies examine neural and environmental risk for attention problems in isolation. Here, we test a model linking a holistic measure of youths’ environment and experiences, the “exposome”, to attention problems via attention-related brain networks. We then test the reciprocal pathway: whether these brain networks are indirectly associated with attention problems via the exposome. This bidirectional framework integrates functional brain organization and environmental context to clarify pathways underlying youth attention problems.

The ability to sustain attention differs among individuals and develops gradually through adolescence (1,2). Impairments of sustained attention are a core characteristic of many psychiatric and neurodevelopmental disorders including attention-deficit/hyperactivity disorder (ADHD), which is associated with negative life outcomes such as worse school performance and increased lifetime risk for substance use, criminality, and suicide (3,4). Clinical evaluations of ADHD rely on multi-informant assessments of attention function as ADHD diagnostic criteria require the occurrence of problems in multiple settings (e.g., at home and in school (5,6)). It is therefore critical to understand how and why attention problems reported by different sources vary among children and change over time.

Many aspects of the physical and psychosocial environment — from pre/perinatal events (7–9), stress (10–15), family cultural values (16–18), household income (19–23), state-level macro-economic factors (24–27), and toxin exposure (19,28–30) — are associated with attentional functioning and brain development, and are thus important candidate mechanisms. Adolescence is a particular period of interest due to the protracted structural development of the prefrontal cortex, hierarchical functional development, and dynamic changes in societal demands and experiences during this timeframe (31–37). Prior work has primarily focused on investigating the effects of environmental factors on development one at a time, for example, examining the role of socioeconomic status and parental education as a proxy for the complexities of everyday experiences (21,36,38,39). However, psychosocial experiences and external environments are interconnected and often have additive and multiplicative effects on an individual’s cognitive and psychiatric outcomes (21,40–44).

To capture these interconnected factors, recent work developed an “exposome” measure encompassing features of an individual’s experiences, exposures, and external environment (41,45–48). The exposome has been associated with obesity (49,50), pubertal stage (49), cognitive task performance (51,52), and psychopathology (48,53–55). The exposome has also been associated with personalized functional topography (51) and functional connectivity (52). Although attention continues to develop throughout adolescence and is shaped by environmental exposures and experiences, how environmental context relates to the neural systems that support attention remains unclear.

Here, we test whether the exposome is associated with caregiver-, teacher-, and youth-reported attention problems and individual differences in attention-related brain network strength. We examine whether the exposome is associated with attention problems both directly and indirectly through these brain networks. Conversely, we test whether predictive brain network strength relates to attention directly and indirectly through the exposome. We hypothesize that environmental exposures and experiences show bidirectional associations with brain network strength which in turn predict subjective reports of attention problems.

## Methods

We tested the associations among multidimensional environmental exposures, an existing fMRI connectome-based predictive model of sustained attention, the saCPM, and attention problems reported from multiple informants. We evaluated the hypotheses that a general exposome factor and an established neuromarker of sustained attention are jointly linked with attention problems, and tested for indirect effects that reflected this shared association.

### Dataset and participants

Data were obtained from the publicly available Adolescent Brain Cognitive Development℠ Study (ABCD Study®) dataset. The ongoing ABCD Study® includes longitudinal annual behavioral and biennial fMRI data from 11,878 youth and their families across 21 sites in the United States (56). Study-wide exclusion criteria at the time of recruitment included standard MRI contraindications, history of traumatic brain injury, birth complications requiring more than 1-month hospitalization, a diagnosis of moderate to severe autism spectrum disorder, intellectual disability, major neurological disorders, schizophrenia, or a substance use disorder (57).

We analyzed behavioral, environmental and resting-state fMRI data from the baseline timepoint, collected when the participants were 9-10 years old. We also included behavioral data from the one-year follow-up (age 10-11) and two-year follow-up (11–12) timepoints. FMRI data were from Release 2.0.1 (http://dx.doi.org/10.15154/1520591). Demographic, environmental and behavioral data were from Release 5.1 (http://dx.doi.org/10.15154/z563-zd24), the most up-to-date behavioral data at the time of analysis. Secondary data analysis was approved by the relevant University of Chicago Institutional Review Board.

### Behavioral measures and covariates

We used data from multiple informants to operationalize attention problems because a key element of diagnosis of Attention Deficit/Hyperactivity Disorder (ADHD) is having attention problems in two or more settings (5). Attention Problem Subscale Ratings were acquired from three harmonized measures: caregiver-report child behavior checklist (CBCL), youth-report brief problem monitor (BPM-Y), and teacher-report brief problem monitor (BPM-T) (58,59). Data were collected when participants were aged 9-10 (baseline), 10-11 (Year 1 follow-up), and 11-12 (Year 2 follow-up) years.

#### Caregiver Reported Attention Problems

Caregivers were asked to report on their youth’s dimensional psychopathology symptoms and behavior using the CBCL at their in-person visit, annually. We used the raw CBCL Attention Problems subscale (CBCL-APS) and controlled for age and sex within each individual model. CBCL-APS scores, *cbcl_scr_syn_attention_r*, are based on 10 of 113 total questions, where caregivers rated their youth on a 0-Not True, 1-Somewhat True, 2-Very True scale.

#### Youth Reported Attention Problems

Youth participants were contacted by phone at least 6-months after their first interview and reported their dimensional psychopathology and general behavior, reflecting on the past 6-month time span. Youth-report attention problems were determined using the BPM-Y attention mean score *bpm_y_ss_attention_mean*, which is a sum of six of 19 individual items on 0-1-2 ratings divided by the total items answered for that subsection.

#### Teacher Reported Attention Problems

At each in-person visit, families were asked to provide the contact information for a teacher with whom the child spent significant time. Teachers were contacted via email and asked to report the behavior of the youth participant, reflecting on a 6-month span. Average teacher-reported attention problems, *bpm_t_ss_attention_mean*, was determined from the same six items as for the BPM-Y.

#### Covariates

Covariates were selected based on factors shown to affect attention in prior literature. Demographics (biological sex and age at the time of interview) and family socioeconomic status (SES) variables (combined household income and caregiver’s level of education) were extracted from *abcd_p_demo.csv*. We included the informant as a covariate, referring to which caregiver was the reporter for all analyses including the caregiver-report CBCL scales, to account for possible caregiver-bias in our sample. Informant *demo_prim* was recorded as either 1: Biological Mother, 2: Biological Father, 3: Adoptive Parent, 4: Child’s Custodial Parent, 5: Other. We include pubertal status because it is associated with both our exposome variable and cognition (49). Perceived Puberty status was extracted from Caregiver-reported “*ph_p_pds.csv*” and Youth-reported “*ph_y_pds.csv*” as calculated Tanner-staging reported on the Pubertal Developmental Scale categorized sum score: Stage 1; pre-puberty, Stage 2; early-puberty, Stage 3; mid-puberty, Stage 4; late-puberty, Stage 5; post-puberty. We selected the first non-missing value from the Pubertal Developmental Scale categorized sum score for youth self-report or caregiver-report survey data, with youth reports prioritized before caregiver reports are used (60,61). Head motion (mean framewise displacement) and number of usable resting-state scans were included as covariates in all imaging analyses. Site and family structure were included as random effects. While race is an important social construct in the United States, our central analysis is not focused on characterizing the features of the environment that are specific to racism, and thus, we will not include race as a covariate in these analyses (62). We evaluated the imaging and behavioral sample to ensure balanced representation across racial categories (**Table 1**).

**Table 1:** Comparison of Behavior-Only to Imaging Sample Characteristics. A chi-squared test was performed to compare the proportion of sex at birth for each group. There is no significant difference between the two samples in sex at birth or age at baseline. A Wilcoxon rank-sum test was conducted to compare the proportions of participants in each race/ethnicity category and pubertal status in the imaging and behavioral samples. A student’s *t*-test was performed to compare the mean of variables of interest and covariates for each sample.

| **TABLE 1: COMPARISON OF BEHAVIOR-ONLY AND BRAIN NETWORK SAMPLE CHARACTERISTICS** |  |  |
| --- | --- | --- |
|  | **Behavior** | **fMRI & Behavior** |
| Number of Participants | 11868 (47.8% Female) | 6691 (48.7% Female) |
| Sites | 21 sites | 19 sites |
| Sex at Birth | $\chi^2 = 7.592 \times 10^{-3}$, N.S. | |
| Male | 6186 | 3431 |
| Female | 5675 | 3258 |
| Age at Baseline | $9.91 \pm 0.625$ | $9.92 \pm 0.63$ |
| Race/ Ethnicity | $W = 17347871$, N.S. | |
| White | 6173 (52%) | 3501 (52%) |
| Black | 1782 (15%) | 958 (14%) |
| Hispanic | 2410 (20%) | 1404 (21%) |
| Asian | 252 (2% ) | 114 (1.7%) |
| Other | 1247 (10.5%) | 713 (10.6%) |
| Reported Pubertal Status | $W = 16823135$, N.S. | |
| Tanner Stage 1 | 3705 | 2053 |

**Table 1: Comparison of Behavior-Only to Imaging Sample Characteristics.**
| **TABLE 1: COMPARISON OF BEHAVIOR-ONLY AND BRAIN NETWORK SAMPLE CHARACTERISTICS** |  |  |  |
| --- | --- | --- | --- |
| Tanner Stage 2 | 4215 | 2421 |  |
| Tanner Stage 3 | 3525 | 1983 |  |
| Tanner Stage 4 | 274 | 155 |  |
| Tanner Stage 5 | 27 | 17 |  |
| Caregiver-report Child Behavior Checklist | 2.98 ± 3.49<br>(N = 10,737) | 2.87 ± 3.42<br>(N = 6,626) | t(10877) = 3.9307, p < 0.0001 |
| Youth-Report Brief Problem Monitor | 0.528 ± 0.423<br>(N = 10,347) | 0.526 ± 0.423<br>(N = 6,371) | t(10578) = 1.8203, N.S. |
| Teacher-Report Brief Problem Monitor | 0.455 ± 0.526<br>(N = 4,644) | 0.4198 ± 0.509<br>(N = 3,066) | t(3843) = 5.5881, p < 0.0001 |
| General Exposome Factor | 0.064 ± 1.045<br>(N = 11,868) | 0.097 ± 1.004<br>(N = 6,691) | t(10618) = -3.8385, p < 0.001 |
| **Exposome Subfactors** |  |  |  |
| School | 0.0147 ± 0.972<br>(N = 11,868) | -0.0212 ± 0.949<br>(N = 6,691) | t(10853) = 4.5717, p < 0.001 |
| Family Values | -0.0893 ± 0.915<br>(N = 11,868) | -0.108 ± 0.910<br>(N = 6,691) | t(11069) = 2.5345, p = 0.011 |
| Family Turmoil | 0.0144 ± 0.94<br>(N = 11,868) | 0.018 ± 0.947<br>(N = 6,691) | t(11213) = -0.48806, p = 0.6255 |
| Dense Urban Poverty | 0.003 ± 1.14<br>(N = 11,868) | -0.112 ± 1.136<br>(N = 6,691) | t(11132) = 12.541, p < 0.001 |
| Extracurriculars | -0.931 ± 0.762<br>(N = 11,868) | -0.915 ± 0.746<br>(N = 6,691) | t(10874) = -2.5371, p = 0.011 |
| Screen Time | -0.506 ± 0.66<br>(N = 11,868) | -0.532 ± 0.653<br>(N = 6,691) | t(11014) = 4.8664, p < 0.001 |

### Functional MRI data, processing, and exclusion criteria

Details of acquisition for functional MRI data for the ABCD Study are described in Casey et al. 2018 (56). We analyzed a subset of minimally processed functional and structural data from the ABCD Study® Curated 2.0.1 (63). Minimal processing included head motion correction, B0 distortion correction using the reversing gradient method, gradient nonlinearity distortion correction, and multi-modal registration using mutual information.

A custom preprocessing pipeline was developed in fMRIprep (64) and detailed in Stier et al 2023 (65) and Kardan et al., 2022 (66). Each structural T1-weighted scan was defaced, skull-stripped, and normalized to the standard MNI152 nonlinear sixth generation template. We utilized the resting-state data, where participants completed four 5-minute runs, up to 20 minutes of eyes-open resting state scans. Each functional run was normalized to T1w space, aligned to MNI space, and then physiological noise regressors were extracted. Framewise displacement was calculated for each functional run using the implementation of Nipype (67). Additional fMRI preprocessing steps include FreeSurfer normalization, ICA-AROMA denoising, CompCor correction and censoring of high motion frames within a 0.9 mm framewise displacement threshold. To further remove motion artifacts, we performed a 36 parameter regression that consisted of the time courses of mean global signal, cerebrospinal fluid signal, white matter signal, the six standard affine motion parameters (x, y, z, pitch, roll, and yaw), their squares, their derivatives, and the squared derivatives of each of these signals (67,68). We used AFNI’s 3dBandpass command to apply a bandpass filter with a high pass cutoff of 0.008 Hz and a low-pass cutoff of 0.12 Hz. Further details of this preprocessing protocol are previously reported (65,66).

We first excluded participants without functional MRI data or who were scanned using Phillips scanners due to a known error in the phase encoding direction while converting from DICOM to NIFTI format, resulting in 9,446 participants from 19 sites being retained. Of those participants, we excluded 100 participants who failed a visual quality check of their T1 structural scan. Next, we applied a frame displacement (FD) threshold of FD mean < 0.5 mm to remove scan runs with excessive head motion, resulting in 6,691 participants with at least one valid resting-state run. Participants with missing data for any variables of interest: teacher-(N=3,066), youth- (N= 6,371), caregiver- (N = 6,626) reported attention problems or covariates: site, family, puberty stage, sex at birth, age at time of interview, parental education and household income were further excluded, resulting in our final sample (**Table 1**).

### Functional connectivity matrix generation

The preprocessed volumetric BOLD images were spatially averaged into the 268-node Shen functional atlas (69). For each participant and run, a whole-brain functional connectivity matrix was calculated by computing the Pearson correlation between the BOLD signal time courses for every pair of nodes and then applying Fisher’s z-transformation, resulting in a 268×268 functional connectivity matrix. Functional connectivity matrices were averaged across all usable runs for each subject, so each participant has one 268×268 matrix. For network-specific analyses, the 268 nodes were clustered into 8 large-scale functional networks defined in previous work (70): medial frontal, frontoparietal, default mode, subcortical-cerebellum, motor, visual I, visual II and visual association.

### Sustained attention network strength calculation

We utilized a previously defined neuromarker, the sustained attention connectome-based predictive model (saCPM; (71)) as an index of sustained attention in our sample. The saCPM was defined to predict sustained attention task performance using fMRI data collected from 25 adults who performed a gradual-onset continuous performance task (71–73). Functional networks that predicted performance included a “high attention” network and a “low attention” network composed of 757 and 630 functional connections, or edges, respectively. Edges in both the high-attention network (predicting better attention function) and low-network (predicting worse attention function) span distributed cortical, subcortical, and cerebellar regions. Mean edge strength in these networks has generalized to predict performance on a stop-signal task (74), zero-back task (66) and clinician-rated ADHD symptoms (71) in out-of-sample populations. The predefined binary high-attention and low-attention masks were downloaded from github.com/monicadrosenberg/Rosenberg_PNAS2020.

saCPM strength was calculated for each participant as the sum of scalar product between all functional connectivity values in the high-attention and low-attention masks, respectively. This sum is linearly related to mean functional connection strength in the networks since there were no missing nodes. Overall saCPM network strength was calculated by subtracting low-attention from high-attention network strength to avoid multicollinearity in regression models.

### Exposome measures

To investigate associations with each child’s multidimensional environmental context, we leveraged a measure of the “exposome” that has been defined previously in the ABCD Study sample (49,51). Details of the exposome definition used in the present study are described in Keller et al. (2024; (51)). The “general exposome” represents the totality of co-occurring features comprising each child’s unique complex experiences and environment. 354 variables were first reduced to data-driven summary scores, and harmonized across multiple formats, lengths, and informant sources (youth-report, caregiver-report, and geo-coded data; **Figure 1A**). The general exposome factor was developed using longitudinal bifactor analysis. Intermediately, 32 sub-scales were determined by a combination of interpretability and subjective evaluation following exploratory factor analysis. Those 32 subscales and additional demographic information were then submitted to longitudinal bifactor modelling, clustered by family and stratified by data collection site. This approach yields equal factor loadings at each timepoint, allowing for longitudinal analyses. The final factors were chosen based on assessments of model fit and interpretability (51), yielding six orthogonal subscales and a general exposome factor.

**Figure 1:**
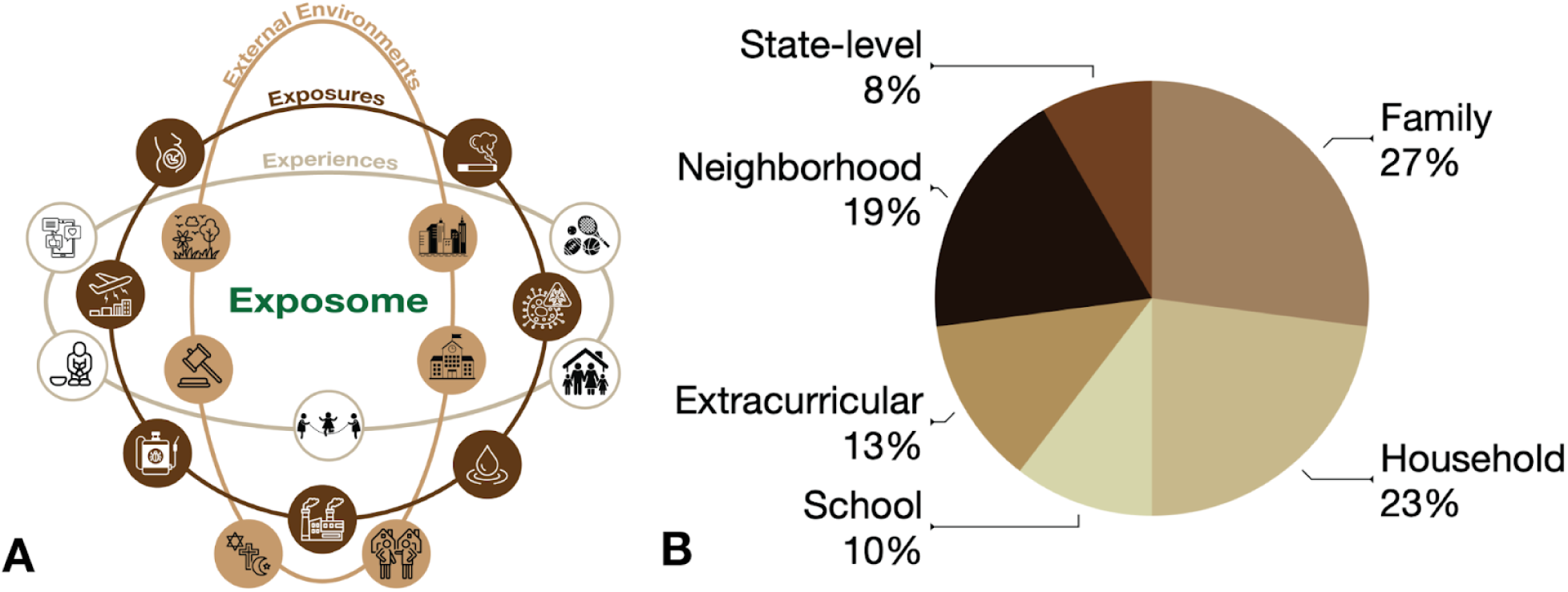
Features of the exposome are multi-level and multi-dimensional. The exposome is a mathematical representation of the totality of external environments, exposures, and experiences through an individual’s lifespan. (**A**) Interlocking rings represent the core domains of an individual’s exposome: physical and chemical exposures (dark-brown ring), social experiences (tan-brown ring), and external environments (caramel-brown ring). An example measurement for each domain is shown within each circle. (**B**) 354 variables from caregiver-report, youth-report, geocoded data were reduced into 32 sub-scales, including parent education, household income, and parental marital status. The pie chart displays the percentage contribution of each multilevel contextual environmental domain to the general exposome factor and six orthogonal sub-factors.

Features such as household income, parental education, and parental marital status loaded positively onto the general exposome factor, whereas factors such as address-based geocoded neighborhood poverty levels, high family religiosity, and low family image loaded negatively on the general exposome factor. Here, we use the general exposome factor detailed in Keller et al. (2024), after multiplying by −1 to align with previous work (46,49,53), such that higher scores reflect greater adversity (e.g., lower household income and higher neighborhood poverty).

To increase the specificity of our analyses, we also examined associations across distinct environmental domains using exposome subfactors. These six previously defined subfactors include school, family values, family turmoil, dense urban poverty, extracurriculars, and screen time (51). These subfactors are necessarily orthogonal to each other and to the general exposome factor. Variables related to grades, school enjoyment and positive feedback at school loaded positively on the school subfactor. Variables captured by the caregiver- and youth-report Mexican American Cultural Values survey — family security, high family image, religiosity and independence — loaded positively on the family values subfactor. Variables related to high family conflict and substance use attitudes positively loaded onto the family turmoil subfactor. Variables related to address-based geocoded neighborhood density, walkability, group living, poverty, air pollution, and ozone loaded positively onto the dense urban poverty subfactor, whereas high levels of neighborhood safety loaded negatively onto this subfactor. The extracurricular subfactor captured the level of involvement a participant had in sports or activities. The final subfactor, screen time, primarily captures the amount of screen time a participant has. The school, extracurriculars, and family values subfactors were multiplied by −1 to conceptually align with the general exposome factor. The dense urban poverty, screentime, and family turmoil subfactors remained the same to preserve the positive relationship between these factors and attention problems.

### Linear mixed effects modeling

Linear regression models were conducted using the lme4 package in R Version 4.5.0 (75). The nuisance covariates include age at the time of interview, sex, informant (for caregiver-report models), and pubertal Tanner Stage. Random effects for family and site were included in all models. To test the association between the general exposome factor and attention problems, we constructed separate mixed-effects models for each attention problem scale as the dependent variable. General exposome factor and the nuisance covariates were modeled as fixed effects with random effects for family and site.

`Caregiver-reported CBCL-APS (raw) ∼ General Exposome Factor + Sex + Age + Puberty + Informant + (1 | Site) + (1 | Family)`

`Teacher-reported BPM-T (mean) ∼ General Exposome Factor + Sex + Age + Puberty + (1 | Site) + (1 | Family)`

`Youth-reported BPM-Y (mean) ∼ General Exposome Factor + Sex + Age + Puberty + (1 | Site) + (1 | Family)`

Next, since the general and specific exposome factors are orthogonal, we included all general and specific subfactors in one model along with the nuisance covariates with random effects for family and site.

Attention problems (caregiver-, teacher-, or youth-report) ∼ General Exposome Factor + School + Family Values + Family Turmoil + Dense Urban Poverty + Extracurriculars + Screen time + Sex + Age + Puberty + Informant + (1 | Site) + (1 | Family)

To test the association between attention problems and saCPM strength, we constructed three separate mixed-effects models predicting different informant reports. We included the number of usable scans and mean framewise displacement as fixed effects.

`Attention problems (caregiver-, teacher-, or youth-report) ∼ saCPM strength + Sex + Head motion + Number of Scans Included + Age + Puberty + Informant + (1 | site) + (1 | family)`

Finally, we tested the association between the general exposome factor and saCPM strength.

`saCPM strength ∼ General Exposome Factor + Sex + Age + Head Motion + Number of Scans Included + Puberty + (1 | Site) + (1 | Family)`

As a sensitivity analysis, we re-ran all of the above models, replacing the general exposome with demographic factors (parental education and household income). We also ran the same models including both the general exposome and demographic factors. This allowed us to evaluate whether the general exposome factor explained variance above and beyond demographic factors, as indicated by changes in conditional R^2^ and Akaike information criterion (AIC) or Bayesian information criterion (BIC).

### Statistical mediation analysis

To test whether the saCPM and exposome operate through a shared pathway, we estimated indirect effects in two complementary statistical mediation models for each attention scale: one with saCPM as the mediator, and the other with the exposome as the mediator. Statistical mediation models were conducted using the mediation package in R with the built-in bootstrapping option for computing *p*-values for the coefficients and the mediated effect (1000 bootstraps) (76). saCPM strength is included as the mediator. Exposome values, adjusted for all fixed effects, are included as the predictor variables, and each questionnaire measure (CBCL, BPMY, BPMT) is modeled as the dependent variable separately. We tested the reciprocal pathway by including the general exposome factor as a mediator. In this analysis, brain-derived saCPM strength, adjusted for all fixed effects, is included as the predictor variable, and each questionnaire measure (CBCL, BPMY, BPMT) is modeled as the dependent variable separately.

### Null model comparison

We tested the specificity of associations between the general exposome factor, attention problem scales, and saCPM by comparing the observed results to null brain network models. We randomly permuted the row and column labels of the square saCPM mask to preserve the mask’s symmetry, calculated null network strength for each participant using this shuffled mask, and related network strength to exposome and attention problem scores with regression models. This procedure was repeated 1000 times. We calculated one-tailed nonparametric *p*-values by comparing observed regression coefficients to a null distribution of coefficients using the following formula:

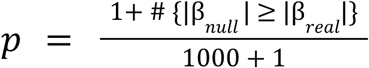

## Results

### The exposome predicts youth attention problems

We assessed cross-informant agreement of attention problems ratings. Highlighting the importance of considering all three informant reports, pairwise correlations were moderate (teacher-report and caregiver report ρ(5038) = 0.467, p <0.001; teacher-report and youth-report ρ(4919) = 0.377, p <0.001; caregiver-report and youth-report ρ(11377) = 0.345, p <0.001).

We performed linear mixed effects modeling to relate attention problems to the exposome. The general exposome factor was associated with more attention problems across all informants (caregiver-report: Std. β = 0.138, SE = 0.01, p <0.001; teacher-report: Std. β = 0.224, SE = 0.02, p <0.001; youth-report: Std. β = 0.215, SE = 0.01, p <0.001; **Figure 2**).

**Figure 2:**
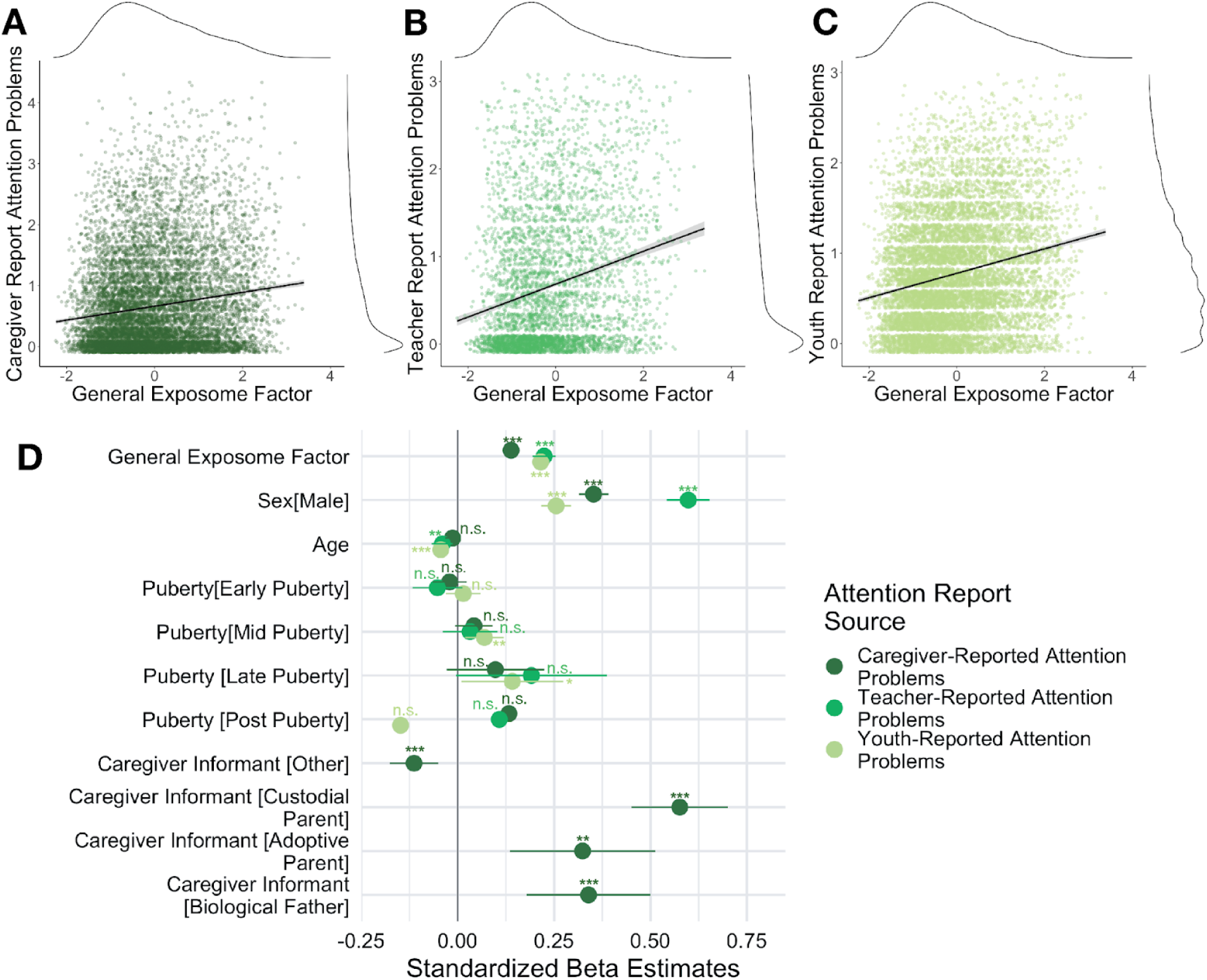
The general exposome factor is significantly associated with caregiver-report (**A**), teacher-report (**B**), and youth-self-report (**C**) attention problems at ages 9-10. Reported attention problems are jittered for visualization. (**D**) The model plot shows fixed-effect estimates from the linear mixed-effects models testing the relationship between the general exposome factor and each cross-informant report. All estimates are shown with 95% confidence intervals and are adjusted for age, sex, pubertal stage, and random effects of site and family. For the models estimating caregiver-reported attention problems, we additionally added the covariate of informant to account for systematic differences in caregiver reporting. Positive coefficients indicate greater reported attention problems associated with higher exposome scores, whereas negative coefficients reflect lower attention problems associated with higher exposome scores.

We tested whether the general exposome factor predicted attention problems over and above the effects of household income and parental education (**Table 2**). We found that the general exposome factor models had a lower AIC and BIC and higher conditional R^2^ values than models only including the demographic variables across all three informants. Models that included both the general exposome factor and demographic variables showed an increase in conditional R^2^, indicating that the general exposome factor contributed additional variance in explaining attention problems, beyond what was explained by the demographic factors alone.

**Table 2:** Model Comparison.

| Table 2: Model Comparison |  |  |  |  |
| --- | --- | --- | --- | --- |
| Informant | Predictor | Marginal R<sup>2</sup>/<br>Conditional R<sup>2</sup> | AIC | BIC |
| Caregiver<br>(N =<br>10735) | General Exposome | **0.063 / 0.342** | **56352.848** | **56471.69** |
|  | SES Variables | 0.057 / 0.334 | 56410.029 | 56626.79 |
|  | General Exposome<br>+ SES Variable | **0.066 / 0.342** | 56354.717 | 56507.62 |
| Youth<br>(N =<br>10345) | General Exposome | 0.068 / 0.298 | **10831.67** | **10911.36** |
|  | SES Variables | 0.050 / 0.286 | 11064.07 | 11230.7 |
|  | General Exposome<br>+ SES Variable | **0.072 / 0.303** | 10853.33 | 10976.48 |
| Teacher<br>(N = 4643) | General Exposome | **0.145 / 0.426** | **6515.795** | **6586.672** |
|  | SES Variables | 0.139 / 0.424 | 6605.003 | 6753.199 |
|  | General Exposome<br>+ SES Variable | **0.146 / 0.426** | 6550.138 | 6659.671 |

#### Exposome subfactors differentially predict attention problems

The general exposome factor and the six sub-factors (school, family values, family turmoil, dense urban poverty, extracurriculars, and screen time) are orthogonal to each other by definition. To test whether dimensions of the exposome differentially predict attention problems, we ran a linear mixed effects model with all 7 variables (general exposome factor and six sub-factors; **Figure S1**). Two of the six sub-factors are robustly associated with attention problems. School adversity (caregiver-report: Std. β = 0.13, SE = 0.01, p <0.001; teacher-report: Std. β = 0.13, SE = 0.01, p <0.001; youth-report: Std. β = 0.23, SE = 0.01, p <0.001) and screen time (caregiver-report: Std. β = 0.05, SE = 0.01, p <0.001; teacher-report: Std. β = 0.10, SE = 0.01, p <0.001; youth-report: Std. β = 0.08, SE = 0.01, p <0.001) both predicted attention problems across all informants.

### Exposome at age 9-10 predicts future attention problems

We investigated longitudinal associations between the general exposome factor and six subfactors at baseline and attention problems one (**Table S1**) and two years later (**Table S2**). The general exposome at baseline predicted teacher-reported and youth-reported attention problems one (**Table S1**) and two years later (**Table S2**), adjusting for attention problems at baseline. Youth with higher environmental adversity had higher reported future attention problems.

Among exposome subfactors, family turmoil predicted attention problems at the 2-year-follow-up session across all informants (caregiver: Std. β = 0.11, p = .001; teacher: Std. β = 0.03, p = .040; youth: Std. β = 0.02, p < .001; **Table S2**). School and family values were associated with higher attention problems in caregiver- and youth-report models at both follow-up visits. Screen time was associated with higher attention problems in the youth-report mode at both follow-up visits.

### A sustained attention neuromarker predicts youth attention problems

As hypothesized, higher saCPM network strength was associated with lower attention problems across all three informants (**Figure 3**; caregiver-report: Std. β = −0.05, SE = 0.01, p <0.001; teacher-report: Std. β = −0.09, SE = 0.02, p <0.001; youth-report: Std. β = −0.07, SE = 0.01, p <0.001). Coefficients were adjusted for head motion, number of rest scans usable, age, sex, and pubertal status with random effects of site and family.

**Figure 3:**
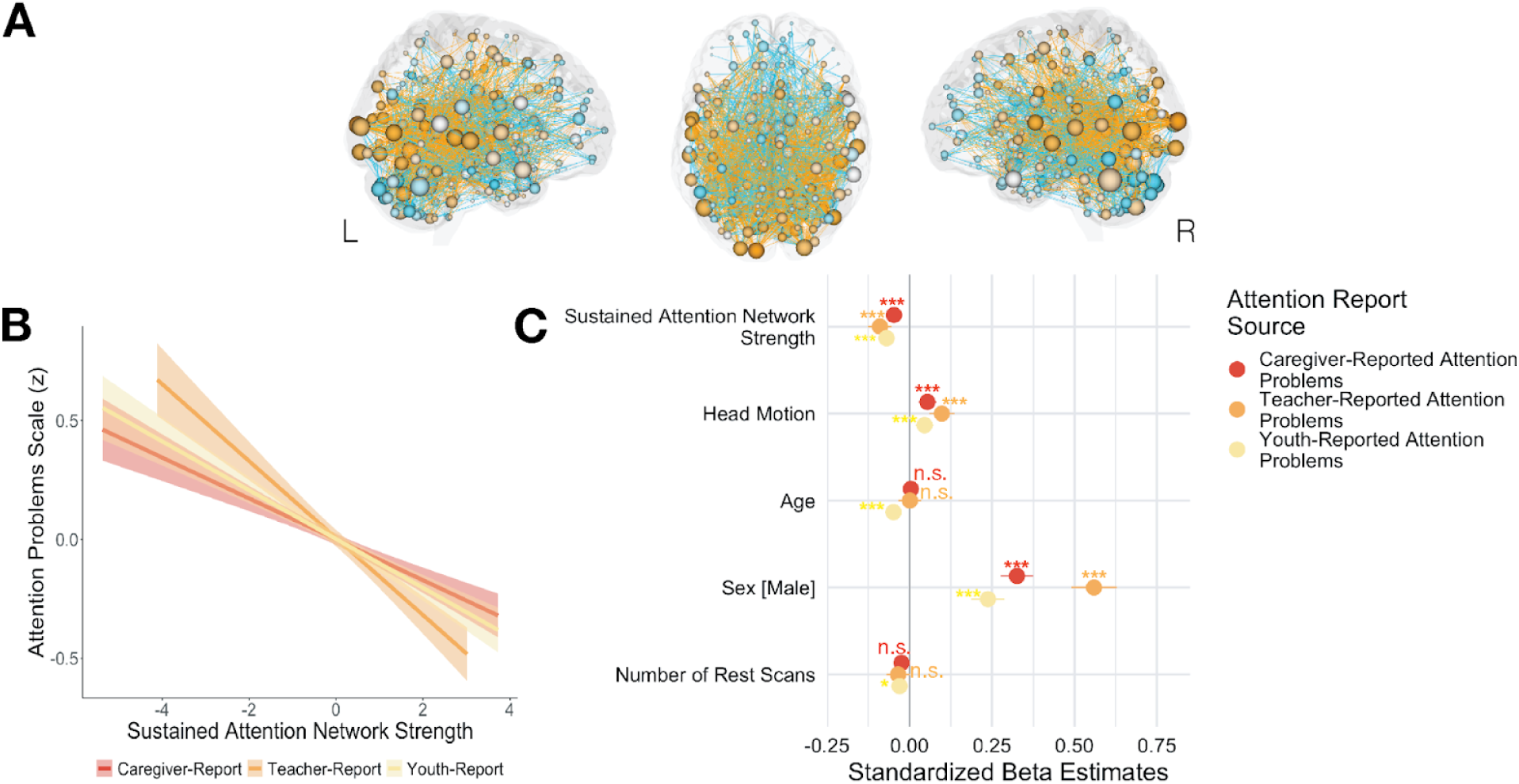
Functional connections in the high-attention (orange) and low-attention (turquoise) networks are significantly associated with attention problems across all three reporters. (**A**) Image adapted with permission from Rosenberg et al., 2017 (72). Nodes are sized according to their total number of attention network connections and colored according to the network in which they have more connections. (**B**) Association between attention problems and saCPM strength from resting-state fMRI data. (**C**) Fixed effects estimates from the three separate linear mixed-effect models testing the relationship between sustained attention network strength and caregiver-report (red), teacher -report (orange), and youth-self-report (yellow) attention problems. All estimates are shown with 95% confidence intervals and are adjusted for head motion, number of usable resting state scans, age, sex, pubertal stage (not pictured), and random effects site and family (not pictured). For the models estimating caregiver-reported attention problems, we additionally added the covariate of informant to account for systematic differences in caregiver reporting.

We tested whether attention problems were more strongly associated with the saCPM’s high- and low-attention networks than with same-size random networks or canonical resting-state networks. Demonstrating specificity, for all three models, beta coefficients for saCPM strength were more extreme than the distribution of null betas for same-sized random networks (two-tailed *p*-value: p<0.001; **Figure 4A**).

**Figure 4:**
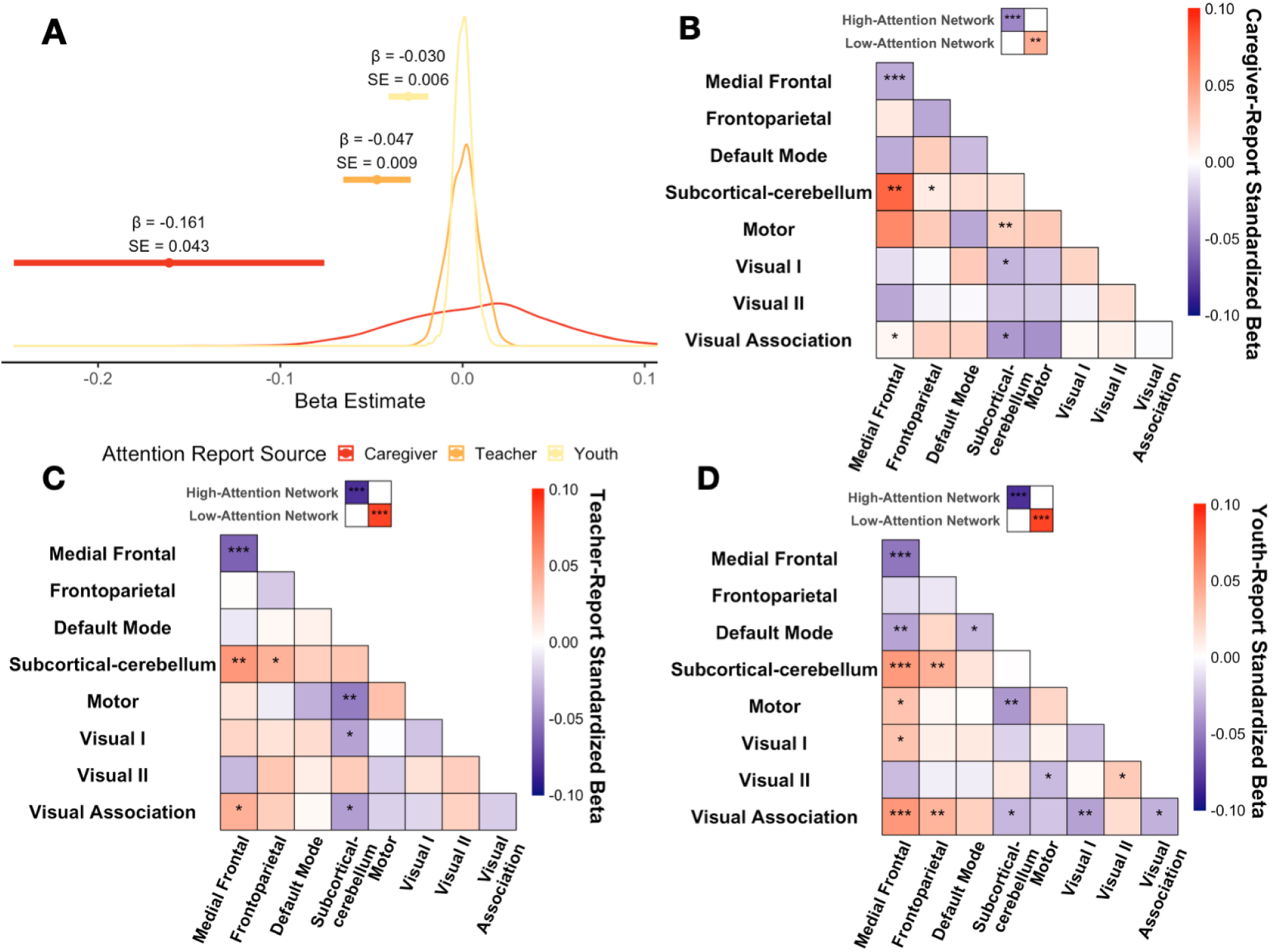
(**A**) The distribution of shuffled null models from 1000 permutations was plotted along with the actual beta estimate with a 95% confidence interval. Associations of the canonical functional networks and caregiver-report (**B**), teacher-report (**C**), and youth self-report (**D**) attention problems. For each canonical network pair, we tested the association between functional connectivity of that network pair and attention problems, adjusted for all covariates.

As expected, when modelled separately, the high-attention saCPM network predicted fewer current attention problems (caregiver-report: Std. β = −0.045, SE = 0.012, p <0.001; teacher-report: Std. β = −0.081, SE = 0.018, p <0.001; youth-report: Std. β = −0.066, SE = 0.013, p <0.001). The low-attention saCPM network predicted more current attention problems (caregiver-report: Std. β = 0.042, SE = 0.013, p <0.001; teacher-report: Std. β = 0.088, SE = 0.019, p <0.001; youth-report: Std. β = 0.064, SE = 0.013, p <0.001). However, baseline saCPM strength was not associated with attention problems at two-year follow-up after accounting for baseline attention problems (**Table S3**).

Functional connectivity within the eight canonical resting-state networks did not outperform the saCPM or the high- or low-attention networks separately in predicting attention problems (**Figure 4**). However, connectivity between the subcortical-cerebellar and medial frontal networks, and between the medial frontal and motor networks, predicted caregiver-reported attention problems better than the low-attention network alone. Across informants, connectivity within the medial frontal network and between the visual association and subcortical-cerebellar networks was associated with fewer attention problems, whereas connectivity between medial frontal and subcortical-cerebellar networks was associated with more attention problems.

### The exposome is associated with functional connectivity signatures of attention

The general exposome factor was associated with saCPM network strength, such that increased environmental adversity corresponded to lower saCPM strength at age 9-10 (Std. β = −0.176, SE = 0.01, p <0.001; **Figure 5A-5B**). This association was stronger than expected from same-sized random networks (two-tailed p < .001; **Figure 5C**), supporting the specificity of the saCPM–exposome relationship.

**Figure 5:**
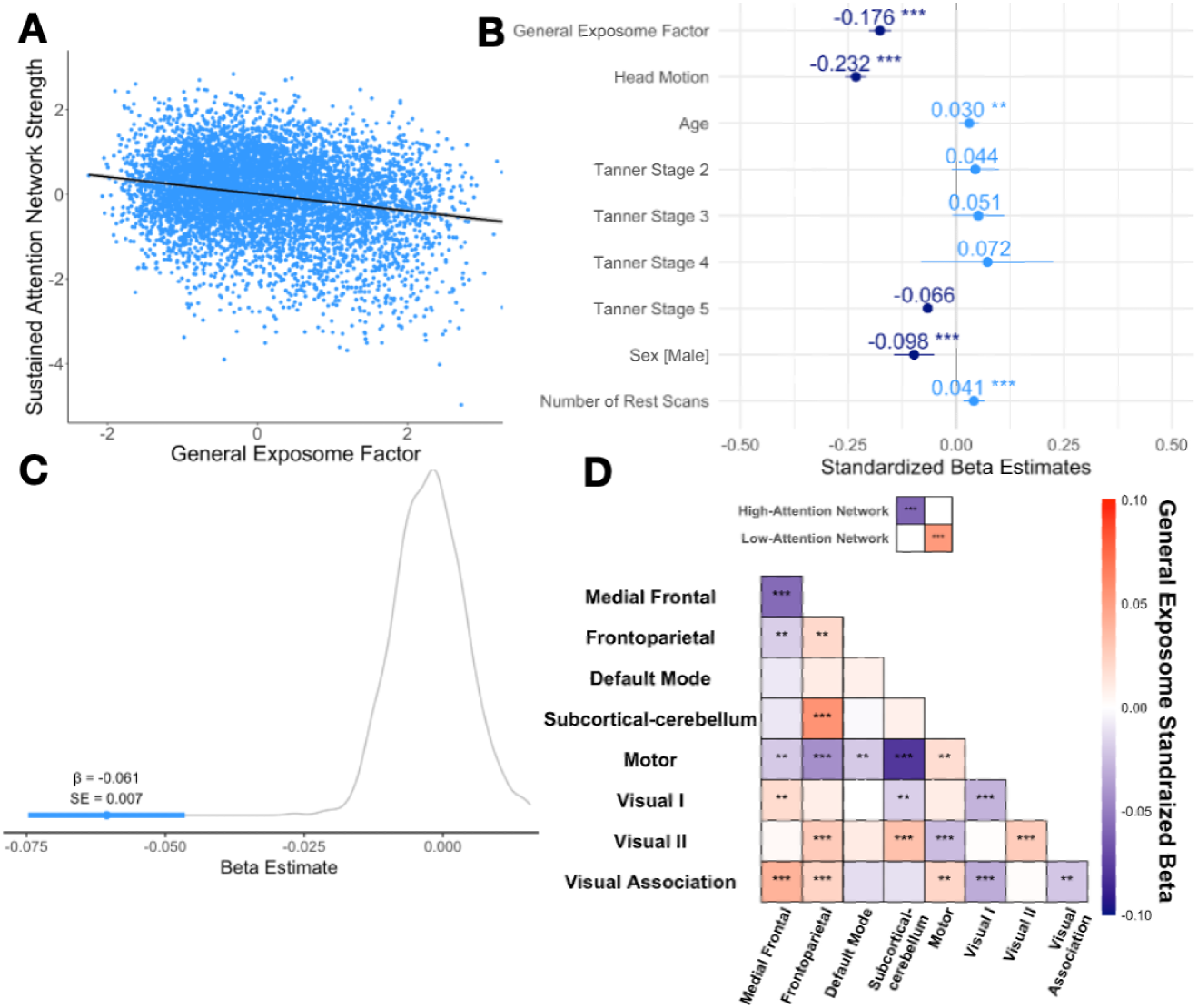
The general exposome factor is associated with sustained attention network strength. (**A**) Association between the general exposome factor and sustained attention network strength, measured using resting-state fMRI. (**B**) Standardized beta estimates predicting sustained attention network strength from a general exposome factor, adjusted for covariates: age, sex, pubertal (Tanner) stage, head motion, and number of rest scans. All estimates are adjusted for random intercepts of site and family and are shown with a 95% confidence interval. (**C**) Distribution of beta estimates from 1000 shuffled null models, along with the actual beta estimate with a 95% confidence interval. (**D**) Associations of the canonical functional networks with the general exposome factor. Statistically significant network pairs are indicated by stars. Network pairs associated with higher general exposome scores are detonated in red; network pairs associated with lower general exposome scores are detonated in blue.

To further assess specificity, we examined associations between the general exposome factor and connectivity within and between canonical resting-state networks (**Figure 5D**).

Lower exposome scores were associated with greater connectivity within the medial frontal, visual I, and visual association networks. Higher exposome scores were associated with greater connectivity within the frontoparietal, motor, and visual II networks. Connectivity between the motor and subcortical-cerebellar networks was more closely associated with exposome scores than the high-attention network alone.

### Indirect associations among the exposome, brain, and attention problems

We tested statistical indirect pathways between the general exposome factor, saCPM, and attention problems at baseline. We examined whether: (1) the saCPM statistically mediated associations between the exposome and attention problems, and (2) the exposome statistically mediated associations between the saCPM and attention problems.

The exposome showed both direct and indirect associations with attention problems reported by all informants. In all three models, the saCPM partially accounted for the relationship between the general exposome factor and attention problems (Caregiver-report ACME Estimate: 0.019, p = 0.008, Proportion Mediated: 4.7%; Teacher-report ACME Estimate: 0.006, p <0.001, Proportion Mediated: 6.1%; Youth-report ACME Estimate: 0.003, p = 0.008, Proportion Mediated: 3.1%; **Figure 6**). Conversely, saCPM strength showed both direct and indirect associations with attention problems. For all three models, the exposome partially accounted for this relationship (Caregiver-report ACME Estimate: −0.014, p < 0.001, Proportion Mediated: 4.7%; Teacher-report ACME Estimate: −0.022, p <0.001, Proportion Mediated: 2.9%; Youth-report ACME Estimate: −0.020, p < 0.001, Proportion Mediated: 4.5%; **Figure 6**).

**Figure 6:**
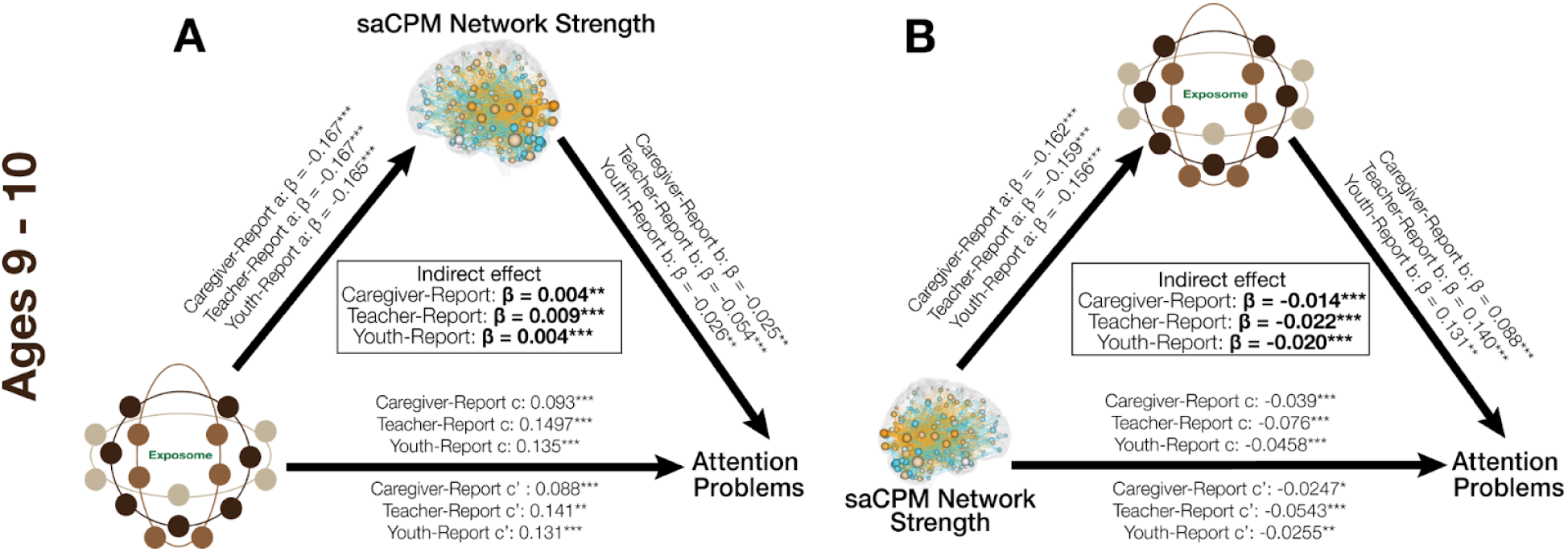
Cross sectional indirect effect models. (A) On the left, saCPM strength as the mediator; (B) panel on the right include the general exposome factors as mediator.

As an exploratory analysis, each exposome subfactor was entered separately as a predictor to examine whether the saCPM network differentially mediated its association with attention problems. We examined these relationships across all three report sources (**Figure S2**). Of the six subfactors tested, the screen time subfactor was the only predictor that showed evidence of significant partial mediation by saCPM network strength across all three report sources. In the caregiver-report and youth-report models, the association between decreased family values subfactor and attention problems showed evidence of partial mediation, such that weaker saCPM network strength accounted for a portion of the association (Caregiver-report indirect effect: Std. β = −0.002, p < 0.01 Std. β = −0.002, p = 0.013; Youth-report indirect effect: Std. β = −0.002, p < 0.01 Std. β = −0.002, p < 0.01).

We explored the role of each canonical network as an indirect pathway between the exposome and attention problems. Across models for different report sources, distinct profiles of canonical networks showed significant indirect effects linking the general exposome factor to attention problems (**Figure S3**).

## Discussion

This study aimed to better understand how environmental exposures and experiences and attention-related brain networks— both on their own and together — relate to attention problems in adolescence. We found that the general exposome factor and exposome subfactors were associated with current and future attention problems, over and above the effects of household income and parent education. The general exposome factor was also associated with the strength of the sustained attention connectome-based predictive model (saCPM), which in turn predicted caregiver-, teacher-, and youth-reported attention problems. Indirect effects analyses indicated that the exposome and saCPM were related to attention problems through partially overlapping pathways. The saCPM partially accounted for the relationship between the exposome and attention problems, and the exposome partially accounted for the relationship between the saCPM and attention problems. These results provide supporting evidence for the bidirectional relationship between the developing brain and an individual’s physical and psychosocial environment.

### saCPM predicts attention problems across reporters

The sustained attention connectome-based predictive model — defined to predict sustained attention task performance in young adults (71) — predicted attention problems in youth rated by all three informants. To our knowledge, this is the first study to test the predictive power of a connectome-based model of cognition for multi-informant subjective reports within the same individuals.

Consistent with assumptions that different informant report sources reflect different behaviors in diverse settings (6), cross-informant agreement for the baseline ABCD Study sample was modest, with moderate correlation between adult reporters (teacher and caregivers) and weak correlation between child-adult pairs (teacher and youth/self report and caregiver and youth/self-report) (59). Despite these differences all three converge on the same attention-related brain network, exceeding what would be expected by chance and by any individual canonical brain network. These results are consistent with the claim that the saCPM network captures a domain-general sustained attention factor (72,77,78).

### The exposome predicts attention problems and attention networks

These results are consistent with the previous literature, showing that multidimensional environments, exposures, and experiences are a key driver of individual differences in attention behaviors (39,51).

Overall, models including general exposome measures were more parsimonious while maintaining equivalent model fit as models including household income and parental education, evidenced by equivalent conditional R^2^ and lower AIC/BIC. The factor loadings underlying the general exposome measures include “typical” socioeconomic variables (i.e., parental education and combined household income) as well as neighborhood safety and census-tracked poverty measures (51); thus, our results add to a large body of evidence that multidimensional socioeconomic context is a strong driver of inter-individual variability in cognitive neurodevelopment (51,79–82).

Our results showed that the general exposome factor is reflected in attention-related FC, such that individuals with increased environmental adversity have decreased attention network strength. Given the breadth of the exposome and the widespread connections in the saCPM network, we might expect varying results for each attention report. However, the results do converge and are consistent with previous work, that the exposome is reflected in functional network organization (51,52) and across different cognitive contexts (47,49,54).

### Exposome subfactors are associated with current and future attention problems

The orthogonal exposome subfactors allow us to understand how varying features of the environment and experiences are independently associated with multi-informant reports. The school and screen-time subfactors of the exposome are robustly related to all three measures of attention, demonstrating that school climate and screen time predict attention behaviors across settings. Our findings corroborate studies investigating the effects of school climate and student-involvement in school as protective factors for mental health and prevention of attention problems (33,83–85). The relationship between screen-time and attention behaviors is well-documented, however, the mechanistic explanation remains unclear (86–88). The family-values and family turmoil subfactors were also robustly related with current and future attention problems, in line with the well-documented consequences of negative family relationships on adolescent cognitive development (18,89,90). Nevertheless, the subfactor analysis highlights aspects of a child’s environment where interventions may be most effectively applied.

### Shared pathways between the exposome and brain

We capitalized on deep phenotyping of participants in the ABCD Study to explore associations between attention problems, external environments (air pollution, state policy), and experiences (extracurricular, screentime). As hypothesized, our findings reveal a bidirectional pattern of indirect effects: the strength of the attention-related network is associated with the link between the exposome and reported attention measures, while the exposome is likewise associated with the relationship between the saCPM and attention problems across all report sources. This highlights the dynamic interplay between environment and experiences, brain network organization and attention functioning, underscoring the importance of considering these factors together rather in isolation.

The extent to which the external environment is reflected in large-scale brain networks and behavior is a key area of study. Previous work has typically focused on the unidirectional impact of the external environment (environment → brain → behavior), with limited consideration of the specific neurobiological phenotypes that shape experiences (brain → environment → behavior) capable of alleviating clinical symptoms and promoting adaptive cognitive development, which in turn feedback on the broader dynamic system (91–93). Our bidirectional framework capitalizes on the fact that some of the psychosocial variables in the exposome (i.e., screentime, extracurricular, peer deviance) may reflect the consequences of neurobiological systems prior to our time period of study.

### Limitations

Although our work highlights the importance of understanding the contextual factors underlying attention, we acknowledge several limitations. First, our findings are from observational data and are primarily cross-sectional and thus do not imply causality. Future work will be critical for testing how trajectories of environmental exposure and experience and attention-related brain networks jointly shape the development of attention problems across adolescence. Second, this sample is non-clinical in nature, with participants generally exhibiting low levels of attention problems across all three reports. Consequently, the restricted range of attention-related difficulties may limit the generalizability of the findings to a clinically enriched population. Finally, the link between social determinants of health, parental education, socioeconomic status, neighborhood factors, and other psychosocial experiences depends on context. Importantly, data collection and recruitment for the ABCD Study was established based on the catchment areas of the 21 study sites, and therefore, do not reflect the national estimates for race, ethnicity or income (57,94–96). Features of the exposome capture co-occurring variables related to social inequality, and in the United States context, these features vary nonrandomly (79,97,98). It is possible that the relationship between the exposome factors and attention problems will look different in a cross-cultural context. The growing availability of large cohort studies like the UK Biobank and GenerationR of the Netherlands, allows the exploration of possible differences in populations with additional protective (universal health care) and risk factors.

## Conclusions

The present study advances our understanding of how multilevel environmental exposures and experiences relate to attention problems during adolescence. Interactions among the exposome and large-scale brain network organization are jointly associated with attention-related behaviors in childhood and adolescence, supporting a bidirectional framework linking the complexities of environmental factors and functional brain organization.

## Data Availability

Data used in the preparation of this article were obtained from the Adolescent Brain Cognitive Development Study (http://abcdstudy.org). Researchers with an approved NIH Brain Development Cohorts (NBDC) Data Use Certification (DUC) may obtain ABCD Study data.

http://dx.doi.org/10.15154/z563-zd24

## Acknowledgements

Data used in the preparation of this article were obtained from the Adolescent Brain Cognitive Development^SM^ (ABCD) Study (https://abcdstudy.org), held in the NIMH Data Archive (NDA). This is a multisite, longitudinal study designed to recruit more than 10,000 children age 9-10 and follow them over 10 years into early adulthood. The ABCD Study® is supported by the National Institutes of Health and additional federal partners under award numbers U01DA041048, U01DA050989, U01DA051016, U01DA041022, U01DA051018, U01DA051037, U01DA050987, U01DA041174, U01DA041106, U01DA041117, U01DA041028, U01DA041134, U01DA050988, U01DA051039, U01DA041156, U01DA041025, U01DA041120, U01DA051038, U01DA041148, U01DA041093, U01DA041089, U24DA041123, U24DA041147. A full list of supporters is available at https://abcdstudy.org/federal-partners.html. A listing of participating sites and a complete listing of the study investigators can be found at https://abcdstudy.org/consortium_members/. ABCD consortium investigators designed and implemented the study and/or provided data but did not necessarily participate in the analysis or writing of this report. This manuscript reflects the views of the authors and may not reflect the opinions or views of the NIH or ABCD consortium investigators. The ABCD data repository grows and changes over time. The ABCD data used in this report came from <u>10.15154/cqdy-5453</u>. DOIs can be found at http://dx.doi.org/10.15154/z563-zd24. Additional support for this work was made possible from NIEHS R01-ES032295 and R01-ES031074.

We would like to thank the NIH ABCD Innovations and Insights Meeting for inspiring this collaboration and the further development of this project. This work was also supported with resources provided by the University of Chicago Research Computing Center.

## Data Availability Statement

Data used in the preparation of this article were obtained from the Adolescent Brain Cognitive Development Study® (http://abcdstudy.org). Researchers with an approved NIH Brain Development Cohorts (NBDC) Data Use Certification (DUC) may obtain ABCD Study data. The analysis code will be publicly available at https://github.com/niaberrian/ABCDExposomeAttention upon publication.

## Funding Sources & Conflicts of Interest

N.B. was supported by NIH T32GM150375 and NIH T32HD007009. A.S.K. was supported by a NARSAD Young Investigator Award from the Brain & Behavior Research Foundation. A.J.S. was supported by NSF grant BCS-2438783. MBG was supported by NSF S&CC 1952050 and a seed grant from the U.Chicago Institute for Climate and Sustainable Growth. O.K. was supported by the National Institute on Drug Abuse K01 DA059598. R.B. was supported by the National Institute of Mental Health. R.B. reports a relationship with Taliaz Health that includes: board membership and equity or stocks. R.B. reports a relationship with Zynerba Pharmaceuticals, Inc that includes: board membership. A.F.C., T.M.M, and M.D.R. have no relevant disclosures or conflicts of interest.

## Notes

### Author Declarations

The study used only openly available human data originally located at https://abcdstudy.org Data used in the preparation of this article were obtained from the Adolescent Brain Cognitive DevelopmentSM (ABCD) Study (https://abcdstudy.org), held in the NIMH Data Archive (NDA). This is a multisite, longitudinal study designed to recruit more than 10,000 children age 9-10 and follow them over 10 years into early adulthood. The ABCD Study is supported by the National Institutes of Health and additional federal partners under award numbers U01DA041048, U01DA050989, U01DA051016, U01DA041022, U01DA051018, U01DA051037, U01DA050987, U01DA041174, U01DA041106, U01DA041117, U01DA041028, U01DA041134, U01DA050988, U01DA051039, U01DA041156, U01DA041025, U01DA041120, U01DA051038, U01DA041148, U01DA041093, U01DA041089, U24DA041123, U24DA041147. A full list of supporters is available at https://abcdstudy.org/federal-partners.html. A listing of participating sites and a complete listing of the study investigators can be found at https://abcdstudy.org/consortium_members/. ABCD consortium investigators designed and implemented the study and/or provided data but did not necessarily participate in the analysis or writing of this report. This manuscript reflects the views of the authors and may not reflect the opinions or views of the NIH or ABCD consortium investigators. The ABCD data repository grows and changes over time. The ABCD data used in this report came from 10.15154/cqdy-5453. DOIs can be found at http://dx.doi.org/10.15154/z563-zd24. Additional support for this work was made possible from NIEHS R01-ES032295 and R01-ES031074.

