## SupplementTables for "The exposome and attention-related brain networks jointly predict attention problems in early adolescence"

### Supplemental Figures and Tables

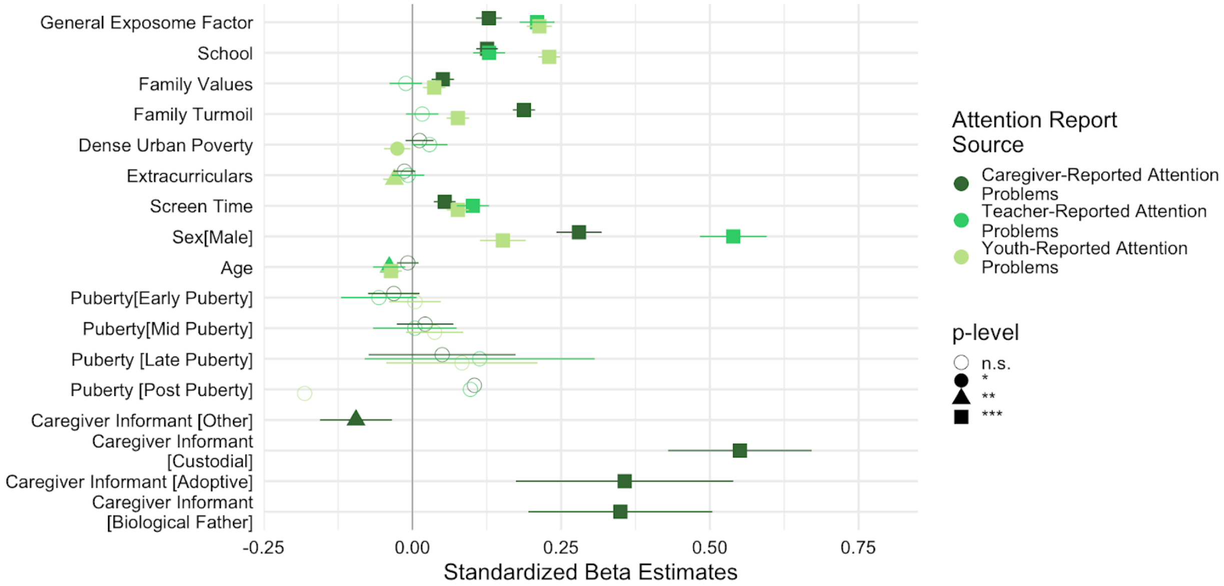


**Figure S1:** Exposome subfactor analysis. The plot shows fixed-effect estimates from linear mixed-effects models testing the relationship between exposome factors and each cross-informant report. For the model estimating caregiver-reported attention problems, we added the covariate of informant. Positive coefficients indicate greater reported attention problems associated with higher exposome subfactors, whereas negative coefficients reflect lower attention problems.

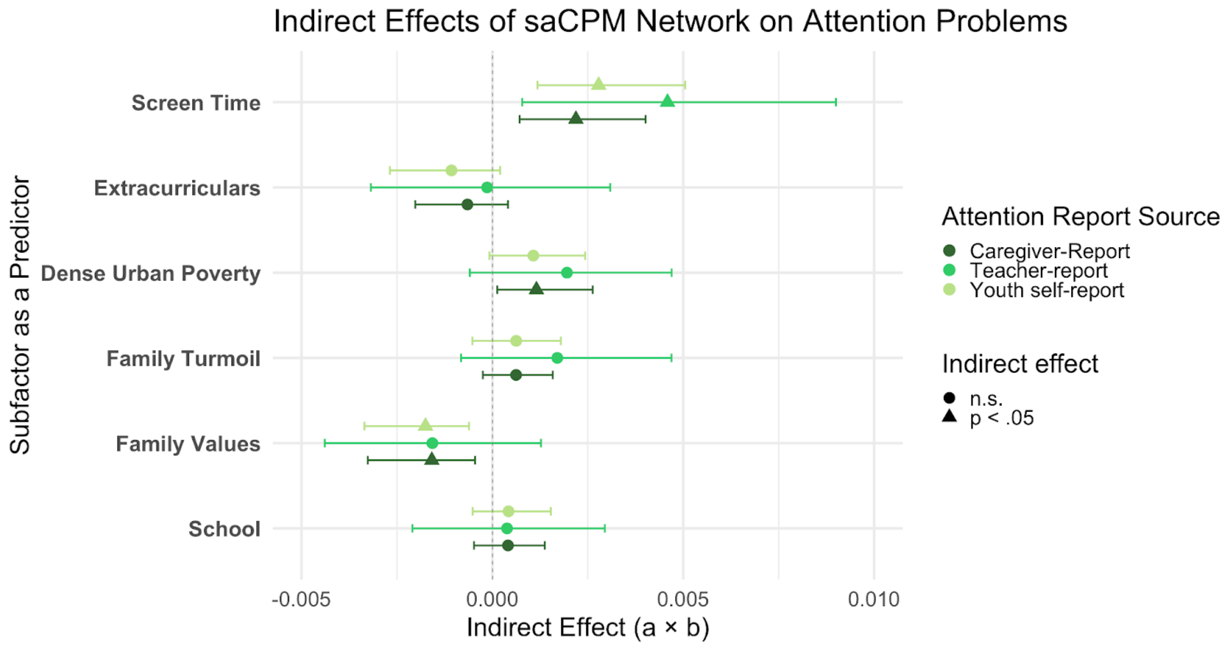


**Figure S2:** Analysis of the indirect effect of functional brain network, saCPM, on the relationship between exposome subfactors and attention measures at age 9-10. Triangles indicate a significant statistical mediation of the saCPM on the relationship between the specific subfactor and each attention measure.

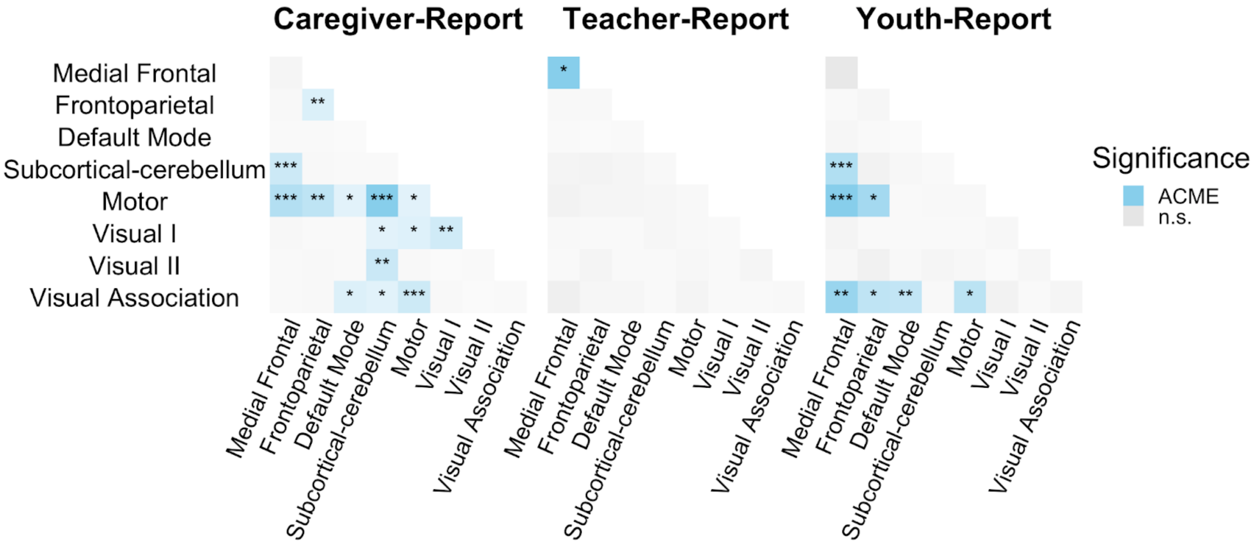


**Figure S3**: Canonical networks that have an indirect effect on the link between the general exposome factor and attention measures across report sources. The transparency of the network reflects the effect size, the darker the color, the larger the indirect effect of the network on the association between the general exposome factor and attention problems.

| | Caregiver-Report Attention Problems at Year 1 | | | Teacher-Report Attention Problems at Year 1 | | | Youth-Report Attention Problems at Year 1 | | |
|---|---|---|---|---|---|---|---|---|---|
| **Variable** | **Beta** | **95% CI** | **p-value** | **Beta** | **95% CI** | **p-value** | **Beta** | **95% CI** | **p-value** |
| **General Exposome Factor at Baseline** | -0.01 | -0.07, 0.05 | 0.7 | 0.06 | 0.04, 0.08 | **<0.001** | 0.03 | 0.02, 0.04 | **<0.001** |
| **School at Baseline** | 0.11 | 0.05, 0.17 | **<0.001** | 0.02 | 0.00, 0.04 | 0.12 | 0.07 | 0.06, 0.08 | **<0.001** |
| **Family Values at Baseline** | 0.09 | 0.03, 0.16 | **0.003** | 0.00 | -0.02, 0.03 | 0.7 | 0.02 | 0.01, 0.03 | **<0.001** |
| **Family Turmoil at Baseline** | 0.06 | 0.00, 0.12 | 0.052 | 0.00 | -0.02, 0.02 | 0.7 | 0.02 | 0.01, 0.03 | **<0.001** |
| **Dense Urban Poverty at Baseline** | 0.02 | -0.04, 0.07 | 0.5 | 0.01 | -0.01, 0.02 | 0.4 | -0.01 | -0.02, 0.00 | 0.3 |
| **Extracurriculars at Baseline** | -0.05 | -0.12, 0.03 | 0.2 | 0.00 | -0.03, 0.02 | 0.8 | 0.01 | -0.01, 0.02 | 0.3 |
| **Screen Time at Baseline** | 0.11 | 0.03, 0.20 | **0.011** | 0.03 | 0.00, 0.06 | 0.086 | 0.04 | 0.03, 0.06 | **<0.001** |
| **Sex at Birth** | | | | | | | | | |
| *Female* | — | — | | — | — | | — | — | |
| *Male* | 0.21 | 0.08, 0.34 | **0.001** | 0.15 | 0.11, 0.19 | **<0.001** | 0.01 | -0.01, 0.03 | 0.2 |
| **CBCL at Baseline** | 0.72 | 0.70, 0.73 | **<0.001** | | | | | | |
| **Age at Baseline** | -0.16 | -0.46, 0.14 | 0.3 | -0.02 | -0.12, 0.08 | 0.7 | 0.00 | -0.05, 0.05 | >0.9 |
| **Age at Year 1 Follow Up** | 0.12 | -0.19, 0.42 | 0.5 | 0.02 | -0.08, 0.13 | 0.7 | 0.00 | -0.05, 0.06 | 0.9 |
| **Pubertal Stage at Year 1 Follow Up** | | | | | | | | | |
| *Tanner Stage 1* | — | — | | — | — | | — | — | |
| *Tanner Stage 2* | 0.00 | -0.15, 0.15 | >0.9 | -0.02 | -0.07, 0.03 | 0.4 | 0.05 | 0.03, 0.08 | **<0.001** |
| *Tanner Stage 3* | -0.06 | -0.22, 0.11 | 0.5 | 0.01 | -0.05, 0.06 | 0.8 | 0.06 | 0.04, 0.09 | **<0.001** |
| *Tanner Stage 4* | -0.03 | -0.30, 0.24 | 0.8 | 0.01 | -0.08, 0.10 | 0.8 | 0.10 | 0.06, 0.15 | **<0.001** |
| *Tanner Stage 5* | 0.58 | -0.60, 1.8 | 0.3 | 0.28 | -0.18, 0.73 | 0.2 | 0.24 | 0.05, 0.43 | **0.015** |
| **Pubertal Stage at Baseline** | | | | | | | | | |
| *Tanner Stage 1* | — | — | | — | — | | — | — | |
| *Tanner Stage 2* | 0.01 | -0.13, 0.15 | 0.9 | 0.01 | -0.04, 0.05 | 0.7 | -0.01 | -0.03, 0.01 | 0.5 |
| *Tanner Stage 3* | 0.00 | -0.15, 0.16 | >0.9 | 0.04 | -0.02, 0.09 | 0.2 | 0.00 | -0.03, 0.02 | 0.7 |
| *Tanner Stage 4* | -0.02 | -0.43, 0.39 | >0.9 | -0.07 | -0.23, 0.08 | 0.4 | -0.03 | -0.10, 0.04 | 0.4 |
| *Tanner Stage 5* | -0.18 | -1.3, 0.90 | 0.7 | 0.58 | 0.14, 1.0 | **0.010** | -0.14 | -0.32, 0.04 | 0.12 |
| **Caregiver Informant at Baseline** | | | | | | | | | |
| *Biological Mother* | — | — | | | | | | | |
| *Biological Father* | 0.30 | 0.04, 0.57 | **0.025** | | | | | | |
| *Adoptive Parent* | 0.62 | -0.45, 1.7 | 0.3 | | | | | | |
| *Custodial Parent* | 0.92 | 0.16, 1.7 | **0.018** | | | | | | |
| *Other* | 0.01 | -0.61, 0.62 | >0.9 | | | | | | |
| **Caregiver Informant at Year 1 Follow Up** | | | | | | | | | |
| *Biological Mother* | — | — | | | | | | | |
| *Biological Father* | -0.50 | -0.76, -0.25 | **<0.001** | | | | | | |
| *Adoptive Parent* | 0.03 | -1.0, 1.1 | >0.9 | | | | | | |
| *Custodial Parent* | -0.56 | -1.3, 0.19 | 0.14 | | | | | | |
| *Other* | 0.15 | -0.50, 0.80 | 0.6 | | | | | | |
| **BPMT at Baseline** | | | | 0.50 | 0.47, 0.54 | **<0.001** | | | |
| **BPMY at Baseline** | | | | | | | 0.53 | 0.51, 0.55 | **<0.001** |

Abbreviation: CI = Confidence Interval

**Table S1:** Linear mixed effects model predicting attention problems at year 1 follow-up from the general exposome factor and the six subfactors, controlling for attention problems at baseline.

| | Caregiver-Report Attention Problems at Year 2 | | | Teacher-Report Attention Problems at Year 2 | | | Youth-Report Attention Problems at Year 2 | | |
|---|---|---|---|---|---|---|---|---|---|
| **Variable** | **Beta** | **95% CI** | **p-value** | **Beta** | **95% CI** | **p-value** | **Beta** | **95% CI** | **p-value** |
| **General Exposome Factor at Baseline** | -0.02 | -0.09, 0.05 | 0.5 | 0.09 | 0.06, 0.12 | **<0.001** | 0.01 | 0.00, 0.02 | **0.045** |
| **School at Baseline** | 0.10 | 0.04, 0.17 | **0.001** | 0.01 | -0.01, 0.04 | 0.3 | 0.07 | 0.05, 0.08 | **<0.001** |
| **Family Values at Baseline** | 0.07 | 0.00, 0.14 | **0.040** | 0.02 | -0.01, 0.05 | 0.2 | 0.01 | 0.00, 0.02 | **0.040** |
| **Family Turmoil at Baseline** | 0.11 | 0.04, 0.17 | **0.001** | 0.03 | 0.00, 0.05 | **0.038** | 0.02 | 0.01, 0.03 | **<0.001** |
| **Dense Urban Poverty at Baseline** | -0.05 | -0.11, 0.02 | 0.14 | 0.01 | -0.01, 0.04 | 0.2 | 0.00 | -0.01, 0.01 | 0.5 |
| **Extracurriculars at Baseline** | -0.03 | -0.11, 0.05 | 0.5 | -0.02 | -0.05, 0.01 | 0.3 | 0.00 | -0.02, 0.01 | 0.6 |
| **Screen Time at Baseline** | 0.07 | -0.02, 0.17 | 0.12 | 0.02 | -0.02, 0.06 | 0.3 | 0.05 | 0.03, 0.06 | **<0.001** |
| **saCPM Network Strength at Baseline** | 0.03 | -0.04, 0.09 | 0.5 | 0.00 | -0.03, 0.02 | 0.8 | -0.01 | -0.02, 0.00 | 0.2 |
| **CBCL at Baseline** | 0.65 | 0.63, 0.67 | **<0.001** | | | | | | |
| **Sex at Birth** | | | | | | | | | |
| *Female* | — | — | | — | — | | — | — | |
| *Male* | 0.23 | 0.08, 0.38 | **0.002** | 0.14 | 0.08, 0.20 | **<0.001** | -0.03 | -0.06, -0.01 | **0.010** |
| **Pubertal Stage at Baseline** | | | | | | | | | |
| *Tanner Stage 1* | — | — | | — | — | | — | — | |
| *Tanner Stage 2* | 0.04 | -0.10, 0.18 | 0.6 | -0.01 | -0.07, 0.04 | 0.7 | 0.02 | -0.01, 0.04 | 0.2 |
| *Tanner Stage 3* | -0.02 | -0.18, 0.13 | 0.8 | 0.00 | -0.07, 0.06 | 0.9 | 0.01 | -0.02, 0.04 | 0.4 |
| *Tanner Stage 4* | -0.32 | -0.73, 0.10 | 0.13 | -0.04 | -0.23, 0.14 | 0.6 | -0.02 | -0.09, 0.05 | 0.6 |
| *Tanner Stage 5* | -0.75 | -1.9, 0.45 | 0.2 | -0.28 | -0.82, 0.26 | 0.3 | -0.11 | -0.31, 0.10 | 0.3 |
| **Pubertal Stage at Year 2 Follow Up** | | | | | | | | | |
| *Tanner Stage 1* | — | — | | — | — | | — | — | |
| *Tanner Stage 2* | 0.06 | -0.14, 0.26 | 0.5 | 0.06 | -0.01, 0.13 | 0.11 | 0.06 | 0.02, 0.09 | **0.001** |
| *Tanner Stage 3* | 0.01 | -0.19, 0.21 | >0.9 | 0.10 | 0.02, 0.18 | **0.015** | 0.08 | 0.05, 0.12 | **<0.001** |
| *Tanner Stage 4* | 0.09 | -0.17, 0.34 | 0.5 | 0.07 | -0.03, 0.17 | 0.2 | 0.09 | 0.04, 0.13 | **<0.001** |
| *Tanner Stage 5* | -0.19 | -0.85, 0.46 | 0.6 | 0.20 | -0.08, 0.49 | 0.2 | 0.16 | 0.05, 0.28 | **0.006** |
| **Caregiver Informant at Baseline** | | | | | | | | | |
| *Biological Mother* | — | — | | | | | — | — | |
| *Biological Father* | 0.34 | 0.06, 0.62 | **0.017** | | | | 0.04 | 0.00, 0.07 | **0.039** |
| *Adoptive Parent* | -2.9 | -4.0, -1.7 | **<0.001** | | | | -0.02 | -0.09, 0.05 | 0.6 |
| *Custodial Parent* | 0.17 | -0.61, 0.95 | 0.7 | | | | -0.01 | -0.12, 0.10 | 0.8 |
| *Other* | -0.04 | -0.68, 0.59 | 0.9 | | | | 0.11 | 0.03, 0.19 | **0.009** |
| **Caregiver Informant at Year 2 Follow Up** | | | | | | | | | |
| *Biological Mother* | — | — | | | | | | | |
| *Biological Father* | -0.59 | -0.86, -0.32 | **<0.001** | | | | | | |
| *Adoptive Parent* | 3.2 | 2.1, 4.3 | **<0.001** | | | | | | |
| *Custodial Parent* | -0.31 | -1.0, 0.39 | 0.4 | | | | | | |
| *Other* | 0.17 | -0.51, 0.84 | 0.6 | | | | | | |
| **Head Motion at Baseline** | 0.05 | -0.01, 0.12 | 0.10 | 0.02 | -0.01, 0.05 | 0.13 | 0.01 | 0.00, 0.02 | 0.12 |
| **Number of Rest Scans at Baseline** | -0.06 | -0.12, 0.00 | 0.064 | -0.01 | -0.03, 0.01 | 0.4 | -0.01 | -0.02, 0.00 | 0.064 |
| **Age at Baseline** | 0.05 | -0.12, 0.21 | 0.6 | 0.00 | -0.07, 0.08 | >0.9 | | | |
| **Age at Year 2 Follow Up** | -0.01 | -0.17, 0.15 | >0.9 | 0.02 | -0.06, 0.09 | 0.7 | 0.04 | 0.02, 0.07 | **0.002** |
| **BPMT at Baseline** | | | | 0.44 | 0.39, 0.49 | **<0.001** | | | |
| **BPMY at Baseline** | | | | | | | 0.45 | 0.43, 0.48 | **<0.001** |
| **Age at time of BPM-Y Baseline** | | | | | | | -0.03 | -0.05, 0.00 | 0.065 |

Abbreviation: CI = Confidence Interval

**Table S2:** Linear mixed effects model predicting attention problems at year-2 follow-up from the general exposome factor and the six subfactors, controlling for attention problems at baseline.

| | CBCL at Year 2 | | | BPMT at Year 2 | | | BPMY at Year 2 | | |
|---|---|---|---|---|---|---|---|---|---|
| **Variable** | **Beta** | **95% CI** | **p-value** | **Beta** | **95% CI** | **p-value** | **Beta** | **95% CI** | **p-value** |
| **saCPM Network Strength at Baseline** | 0.02 | -0.04, 0.09 | 0.5 | -0.02 | -0.04, 0.01 | 0.3 | -0.01 | -0.02, 0.00 | 0.10 |
| **Head Motion at Baseline** | 0.06 | 0.00, 0.13 | 0.059 | 0.03 | 0.01, 0.06 | **0.017** | 0.01 | 0.00, 0.02 | 0.057 |
| **Number of Rest Scans at Baseline** | -0.06 | -0.12, 0.00 | 0.058 | -0.01 | -0.03, 0.01 | 0.5 | -0.01 | -0.02, 0.00 | 0.075 |
| **CBCL at Baseline** | 0.66 | 0.65, 0.68 | **<0.001** | | | | | | |
| **Age at Baseline** | 0.04 | -0.12, 0.20 | 0.6 | 0.03 | -0.05, 0.10 | 0.5 | -0.02 | -0.05, 0.00 | 0.11 |
| **Caregiver Informant at Baseline** | | | | | | | | | |
| *Biological Mother* | — | — | | | | | | | |
| *Biological Father* | 0.35 | 0.07, 0.63 | **0.014** | | | | | | |
| *Adoptive Parent* | -2.8 | -4.0, -1.7 | **<0.001** | | | | | | |
| *Custodial Parent* | 0.14 | -0.64, 0.92 | 0.7 | | | | | | |
| *Other* | 0.02 | -0.62, 0.65 | >0.9 | | | | | | |
| **Caregiver Informant at Year 2 Follow Up** | | | | | | | | | |
| *Biological Mother* | — | — | | | | | | | |
| *Biological Father* | -0.58 | -0.85, -0.31 | **<0.001** | | | | | | |
| *Adoptive Parent* | 3.2 | 2.1, 4.3 | **<0.001** | | | | | | |
| *Custodial Parent* | -0.32 | -1.0, 0.38 | 0.4 | | | | | | |
| *Other* | 0.10 | -0.57, 0.78 | 0.8 | | | | | | |
| **Age at Year 2 Follow Up** | 0.02 | -0.14, 0.19 | 0.8 | 0.00 | -0.08, 0.07 | >0.9 | 0.04 | 0.01, 0.07 | **0.007** |
| **Pubertal Stage at Baseline** | | | | | | | | | |
| *Tanner Stage 1* | — | — | | — | — | | — | — | |
| *Tanner Stage 2* | 0.08 | -0.06, 0.22 | 0.3 | 0.01 | -0.05, 0.06 | 0.7 | 0.02 | 0.00, 0.05 | 0.091 |
| *Tanner Stage 3* | -0.02 | -0.18, 0.14 | 0.8 | 0.02 | -0.04, 0.09 | 0.5 | 0.02 | -0.01, 0.05 | 0.13 |
| *Tanner Stage 4* | -0.30 | -0.71, 0.12 | 0.2 | 0.04 | -0.15, 0.22 | 0.7 | 0.00 | -0.07, 0.08 | >0.9 |
| *Tanner Stage 5* | -0.78 | -2.0, 0.42 | 0.2 | -0.12 | -0.67, 0.43 | 0.7 | -0.08 | -0.29, 0.13 | 0.4 |
| **Pubertal Stage at Year 2 Follow Up** | | | | | | | | | |
| *Tanner Stage 1* | — | — | | — | — | | — | — | |
| *Tanner Stage 2* | 0.04 | -0.16, 0.23 | 0.7 | 0.05 | -0.03, 0.12 | 0.2 | 0.05 | 0.02, 0.09 | **0.003** |
| *Tanner Stage 3* | -0.12 | -0.31, 0.07 | 0.2 | 0.02 | -0.05, 0.10 | 0.5 | 0.09 | 0.05, 0.12 | **<0.001** |
| *Tanner Stage 4* | -0.15 | -0.37, 0.07 | 0.2 | -0.02 | -0.11, 0.07 | 0.6 | 0.10 | 0.06, 0.14 | **<0.001** |
| *Tanner Stage 5* | -0.45 | -1.1, 0.19 | 0.2 | 0.10 | -0.19, 0.38 | 0.5 | 0.17 | 0.06, 0.29 | **0.004** |
| **BPMT at Baseline** | | | | 0.50 | 0.45, 0.55 | **<0.001** | | | |
| **BPMY at Baseline** | | | | | | | 0.50 | 0.48, 0.53 | **<0.001** |

Abbreviation: CI = Confidence Interval

**Table S3:** Linear mixed effects model predicting attention problems at year 2 follow up (ages 11-12) from the saCPM strength at baseline (age 9-10) controlling for attention problems at baseline. saCPM network strength at baseline does not predict attention problems at Year 2 Follow Up, controlling for baseline attention problems.
